# Weight management needs in under-resourced communities elicited using storyboarding and a realist lens: A qualitative study

**DOI:** 10.64898/2026.02.13.26346241

**Authors:** Tracey J Brown, Kate Mahoney, Felix Naughton, Natalie An Qi Tham, Zarnie Khadjesari

## Abstract

**Background:** Overweight and obesity are causing growing public health, economic and clinical burden, particularly within under-resourced communities. There is an urgent need to develop an in-depth understanding of experiences of weight management, and preferences for support within under-resourced communities, with a view to developing more effective weight management interventions.

**Methods:** Focus groups were run in under-resourced communities using storyboarding; a method to facilitate inclusive communication (n=37). Thematic analysis was applied to textual and visual data, and a realist ‘lens’ applied to provide in-depth insight into weight management experiences and needs. We believe this is the first study to use this combined methodology to explore weight management experiences and needs.

**Results:** Combining storyboarding with a realist lens, generated four themes. Living circumstances indicated that mental health, individual needs, and cost of weight management services were key contextual factors. *Mechanisms of weight management* identified emotional eating and portion control to be central to individual weight management. *Yo-yo dieting* centred on participants’ experiences of weight regain after attempting weight loss. *Weight management intervention needs* indicated psychological support was perceived as severely lacking, and the only route to attain sustained weight management. Offering both in-person and online support for weight management was considered important to reach more people.

**Conclusion:** Moving weight management support from short- to long-term and incorporating more robust psychological support would better serve the needs of people living in under-resourced communities who are overweight or obese. Ideally interventions should be multicomponent and tailored to individual needs and circumstances.

## Background

Overweight and obesity are strongly linked to risk of developing non-communicable diseases including cancer, coronary heart disease and diabetes [1]. The global prevalence of overweight and obesity is significant, at an estimated level of 37%, and this level is predicted to rise [2]. As such, overweight and obesity is causing an increasing public health, clinical and economic burden [1,3]. By 2035, obesity is predicted to be the leading modifiable risk factor for non-communicable diseases [1]. The estimated cost of obesity in 2019 was equivalent to 1.8% of gross domestic product, and this is projected to rise to 3.6% by 2060 [3]. Furthermore, there are disparities in obesity prevalence, with a greater burden experienced in marginalised areas due to under-resourcing limiting both individual opportunities and service provision [4]. In under-resourced communities, there is a lower likelihood of uptake of weight management interventions where these exist, due to factors including employment, transport and caregiving conflicts [4,5]. Obesity is a complex problem and requires in-depth understanding to develop solutions, particularly within under-resourced communities [6]. There are relatively few studies which have detailed lived experiences and weight management needs of people with obesity. A qualitative synthesis of studies exploring experiences of people living with obesity found that daily living circumstances and stigma were related to difficulties in accessing weight management services [7]. However, there is a need for further research to understand experiences and service opportunities to facilitate long-term weight management, particularly within under-resourced communities [6,7].

There is a growing call for participatory research in under-resourced communities to understand complex needs and experiences, with the aim of increasing the effectiveness of interventions to promote equitable health in these populations [7,8]. For participatory research to work most effectively, techniques should be applied to increase engagement.

Visual qualitative techniques offer a way to increase engagement by offering participants alternative ways to express themselves [9]. Storyboarding, originating from the film-making industry, is an innovative and creative visual method used as an inclusive approach to elicit rich participant insight [10–13]. Storyboarding typically uses a sheet of paper divided into sections representing a sequential narrative, such as moving from views before, to during and after an intervention, and invites participants to complete these sections using a variety of materials to enable expression through drawing, collaging, and/ or writing [10].

Storyboarding is particularly suited to exploring emotive topics [11,12], such as weight management. In addition to methods to engage communities to understand needs, there is a growing recognition of the importance in understanding complexity to develop effective interventions for multiplex problems, such as obesity. Realist evaluation is increasingly applied to uncover key contextual and mechanistic factors that lead to particular outcomes, and uncover the elements that are needed for successful interventions [14].

We aimed to understand experiences of weight management and preferences for support in under-resourced communities using a novel combination of storyboarding and analysis using a realist ‘lens’. To our knowledge this is the first study to use this combined approach to explore weight management service needs.

## Methods

Our novel approach used storyboard focus groups and a realist ‘lens’ to understand views of weight management services. By combining storyboards with a realist ‘lens’ we aimed to generate rich insight into the complex underlying thought processes of weight management needs and experiences. Ethical approval was granted by the FMH Research Ethics Committee at the University of East Anglia and we followed the Standards for Reporting Qualitative Research (SRQR) (supplementary file 1).

Focus groups were run by two researchers trained to use storyboarding to support visual and inclusive communication. Researchers were academics based outside of the under-resourced communities participating in the research, which may have influenced participant responses. Researchers took the time at the start of each focus group to build rapport and trust. The storyboard approach was also designed to minimise any perceived power imbalance. Further, the storyboard template and topic guide were developed in conjunction with patient and public representatives. An informal meeting was held with patient and public representatives to trial the storyboard exercise. Amendments were incorporated based on their feedback, which centred on reducing the number of activities and increasing the focus group duration.

### Participant recruitment

Focus groups were conducted in community settings in the counties of Essex, Suffolk and Norfolk, UK. To reach under-resourced communities, areas amongst the top 20% most deprived were targeted, based on Index of Multiple Deprivation [15]. Adults (≥18 years) with a self-reported BMI ≥25kg/m^2^, and/or self-identifying as overweight or interested in receiving weight management support were included. We aimed to recruit six groups of 6-8 participants, seeking to cease recruitment once a sufficiently ‘data-rich’ sample had been reached [16].

To identify participants in under-resourced communities we leveraged existing trusted networks and community channels. To reach a diverse range of participants, recruitment flyers, posters, electronic advertisements and email were used to facilitate in-person and remote recruitment. Community group leads aided participant recruitment where needed. Participants were compensated with a £50 voucher, and up to £10 travel reimbursement for their time. Participants expressing an interest were supplied with a user-friendly demographics form and a short, user-friendly version of the full participant information sheet, designed in conjunction with patient and public representatives and community researchers with experience of developing accessible study information (see Cherish research study [17]). Consent was taken formally at the start of the focus groups.

### Data collection

Focus groups were conducted between February and April 2025. The semi-structured topic guide explored views and experiences of personal weight management, weight management services, and future service needs in under-resourced communities. The storyboard activity invited participants to complete an exercise sheet to break down their thoughts before, during and after using a weight management service (supplementary file 2). The storyboard exercise sheet prompted participants to consider contextual lifestyle factors, their feelings, barriers and enablers for their individual weight management needs, and what might encourage them to use a weight management service. As such, the guide was designed to elicit context-mechanism-outcome configurations in line with the realist approach [14]. Participants could complete the sheets by collaging, drawing, or adding a few words about their feelings, and barriers and enablers [10]. Participants were encouraged to present and describe their storyboard to the rest of the group. This enabled researchers to understand meanings behind the images participants had included or drawn on their storyboards. Focus groups were audio-recorded and transcribed verbatim. Data was pseudonymised by replacing names with a code to protect participant anonymity. Photographs were taken of the storyboards at the end of the session and participants were offered the chance to take their storyboard home if they wished to keep it.

### Data analysis

A combination of both textual and visual analysis was applied, facilitated using NVivo version 14. Thematic analysis [18] was used to find key themes, similarities and differences between participant reflections. Data was analysed deductively, guided by questions in the topic guide, and inductively to identify unprompted themes. Transcripts were read and re-read, and codes were assigned to data segments to capture their core meaning. To analyse visual content, storyboards were viewed and re-viewed to enable data familiarisation. A broad description was given to the overall storyboard to include the type of images selected and their positioning and use of colour to aid familiarisation and data interpretation. The specific meaning of the images was interpreted based on the context and description given by the participant [10]. Codes were assigned to image segments to capture core meaning. A realist ‘lens’ was applied to the data by using codes for each context, mechanism and outcome present within transcripts and storyboards, to increase understanding of participant beliefs and offer explanations of phenomena [19]. Codes from both transcripts and storyboards were organised into groups to generate overall themes. Analysis was conducted by the lead author, TJB. A summary of the findings was shared with patient and public representatives who ‘sense checked’ the outcomes and summarised views using a group storyboard which was used to triangulate findings to enhance the credibility of analysis. A summary of the overall findings was additionally shared with participants for information and any feedback.

## Results

Six focus groups were conducted in the community with a total of 37 participants taking part and completing storyboards. Session duration ranged from 82 to 95 minutes. All participants reported an interest in weight management, and all but one participant self-reported as overweight or obese, with a mean BMI of 34.9 (SD 6.77). Full participant characteristics are shown in Table 1. Groups were conducted in community settings across Norfolk, Suffolk and Essex in: Gainsborough; Great Yarmouth; Greenstead in Colchester; Jaywick; Norwich; and Whitton, all of which include some of the 20% most deprived areas in England [20].

**Table 1.** participant characteristics.

| **Characteristics** | **Total n=37** |
| --- | --- |
| **Gender, n (%)** |  |
| Female | 24 (64.86) |
| Male | 13 (35.14) |
| **Age (years), mean (SD)** | 56.35 (11.02) |
| 18-25, n (%) | 0 (0.00) |
| 26-35, n (%) | 1 (2.70) |
| 36-45, n (%) | 5 (13.51) |
| 46-55, n (%) | 12 (32.43) |
| 56-65, n (%) | 12 (32.43) |
| 66-75, n (%) | 6 (16.22) |
| >75, n (%) | 1 (2.70) |
| **Ethnicity, n (%)** |  |
| White British or Irish | 33 (89.19) |
| Asian or Asian British | 2 (5.41) |
| Black or Black British | 0 (0.00) |
| Mixed ethnic groups | 1 (2.70) |
| Other ethnic groups | 1 (2.70) |
| **Employment, n (%)** |  |
| Full-time employee | 6 (16.22) |
| Part-time employee | 9 (24.32) |
| Self-employed | 0 (0.00) |
| Contractor | 0 (0.00) |
| Worker or casual employment | 0 (0.00) |
| Retired | 12 (32.43) |
| Long-term sick or disability | 7 (18.92) |
| Volunteer | 1 (2.70) |
| Unemployed | 2 (5.41) |
| **Qualifications, n (%)** |  |
| Higher education qualification | 16 (43.24) |
| A level or equivalent | 10 (27.03) |
| GCSEs or equivalent | 6 (16.22) |
| None or not specified | 5 (13.51) |
| **Index of multiple deprivation (decile)\*, n (%)** |  |
| 1 | 12 (32.43) |
| 2 | 3 (8.11) |
| 3 | 3 (8.11) |
| 4 | 2 (5.41) |
| 5 | 4 (10.81) |
| 6 | 3 (8.11) |
| 7 | 2 (5.41) |
| 8 | 2 (5.41) |
| 9 | 3 (8.11) |
| 10 | 3 (8.11) |
| **BMI (kg/m<sup>2</sup>), mean (SD)** | 34.97 (6.77) |
| <18.5 (underweight), n (%) | 0 (0.00) |
| 18.5-24.9 (healthy weight), n (%) | 1 (2.70) |
| 25-29.9 (overweight), n (%) | 9 (24.32) |
| 30-39.9 (obese), n (%) | 21 (56.76) |
| ≥40 (severely obese), n (%) | 6 (16.22) |

\* Where Decile 1 is the 10% most deprived, and Decile 10 is the 10% least deprived.

Four main themes were generated guided by a ‘realist lens’ encompassing key contextual circumstances influencing weight and weight management, mechanisms of change, weight management outcomes, and weight management needs. We focus on experiences of participants, supplemented with insights from patient and public representatives.

### Living circumstances

A strong theme related to difficulties in accessing weight management services, or facilities that would support weight management, such as sports facilities. Participants reported a lack of local services and limitations in transport to these services. Some participants referred to feelings that their local neighbourhood was ‘going downhill’ and a sense of loss of community spirit due to lack of resources and closure of some services:

> *‘when that [community centre] was open they used to do lots of stuff down there, cooking and all different, but that shut down, didn’t it?’* (participant 005)

Other participants spoke positively of local initiatives to improve wellbeing but there were some reports that these services were not well attended, indicating barriers beyond simple locality of services. Participants repeatedly referred to barriers of low income and high costs of accessing healthier foods, physical activities, and weight management services:

> *‘can’t afford to eat expensive, healthy food…I think that’s the big thing with weight management, it is easy to fill up cheaply’* (participant 019)

> *‘Anything with weight attached or diet attached, money making. They’re not overly there to help you I feel*.*’* (participant 020).

Participants spoke about the context of culture and an increasing access to less healthy foods and more sedentary lifestyles, which was described by one participant as a society ‘drowning in obesity’. In part, this stemmed from historical cultural eating practices, giving ‘love through food’, perpetuating less healthy eating preferences. It was acknowledged that healthcare resources could only reach so far to rectify this culture and that individuals also played a role in their behaviours. Participants commonly selected images of less healthy foods and images of sitting down to illustrate their perceptions of their lifestyle before a weight management intervention (Figure 1).

**Figure 1.**
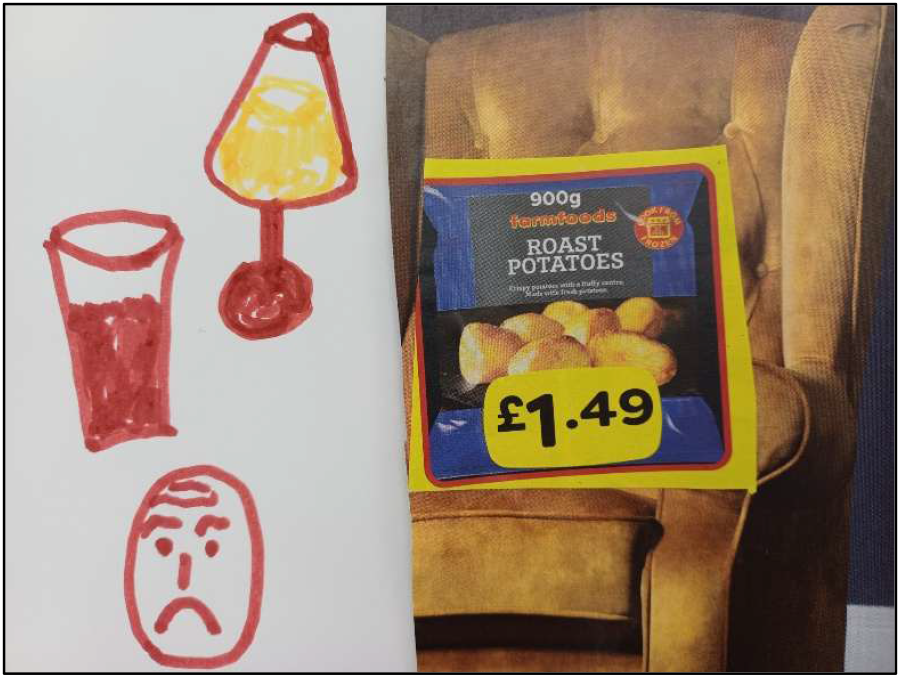
Storyboard extract, participant 015

However, there were significant barriers to initiating positive behavioural changes due to medical conditions, disabilities, mental health needs and living circumstances. Co-morbidities frequently contributed to weight gain and also resulted in barriers to physical activity and undertaking daily activities:

> *‘Large doses of steroids three times a day, and that’s making me put on weight. That makes me put on weight, a lot of weight, and that’s really hard to deal with. And that’s also a problem when it becomes going to the gym, because if I overwork myself I end up having an…attack, because I can’t control my body heat. So I can’t do too much exercise either. So that is a big part of my weight problem is because I’m on steroids, still on steroids three times a day. And then obviously probably my food is not helping on top of that*.*’* (participant 002)

Some participants represented thoughts of personal barriers on their storyboards using sad or anxious faces to illustrate their feelings (Figure 2).

**Figure 2.**
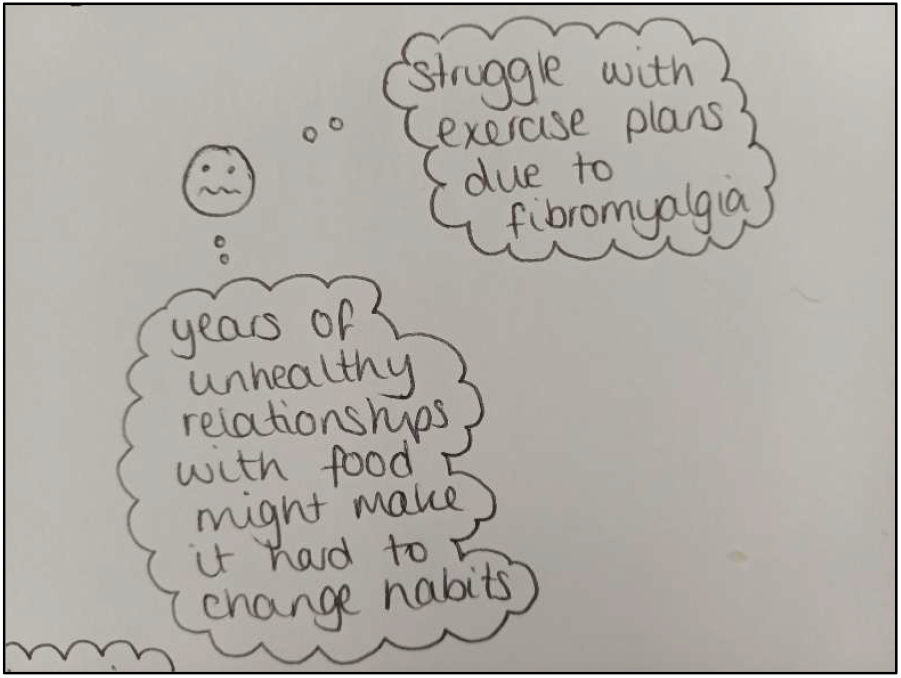
Storyboard extract, participant 033

Participants spoke about mental health challenges and how mental health contributed to weight gain due to mental health medication, food ‘addiction’, and by limiting opportunities for physical activity and getting out into the community. Participants expressed strong feelings of frustration in lack of support and advice for weight maintenance in the context of co-morbidities, mental health, and individual needs:

> Participant 021
>
> *‘overweight is an addiction ‘cause you’re addicted to food! There’s no help out there. No help out there. You can get help for alcohol, drug use –*
>
> Various
>
> *Yeah*.
>
> Participant 023
>
> *But eating a lot you can’t*.
>
> Participant 021
>
> *And it costs us to go somewhere to try and lose it*.*’*

A number of participants referred to chronic life changes which acted as barriers, including an increase in weight due to aging, a gradual reduction in capacity to exercise; or specific to women, the menopause. Acute life events could also act as significant barriers, including deeply personal traumatic events from childhood or adulthood. The most commonly reported chronic life events were illness and physical incapacity reducing activities and causing medication related weight gain. Becoming a parent was also raised, with participants reporting barriers to weight management due to lack of time and prioritising children. Lack of time due to working circumstances was additionally raised as a limiting factor. Living alone, isolation or lack of a partner were also perceived as barriers to eating healthily:

> *‘I just don’t see the point. Living on your own you’re just cooking for one’* (participant 010)

Where life events were attributed to weight-related factors, this could prompt participants into action to make behavioural changes and to manage their weight:

> *‘it took me to have four heart attacks before I realised that’s what I needed to do*.*’*

(participant 003)

**Mechanisms of weight management**

A variety of living circumstances triggered a variety of different emotional responses which could motivate or demotivate participants to engage with weight management support or to make individual changes. A strong theme was the influence of negative thoughts, including anxiety, stress, depression and fatigue, which prompted emotional eating:

> *‘I just love food! And I find it gives me a lift and it gives me a comfort and it gives me a friend, it gives me a companion. And it’s terrible to say that, I know, but that’s the truth*.*’* (participant 008)

Participants represented the influence of negative thoughts and emotional eating on their storyboards by selecting or drawing images of despondent people and using images of ‘treat’ foods (Figure 3).

**Figure 3.**
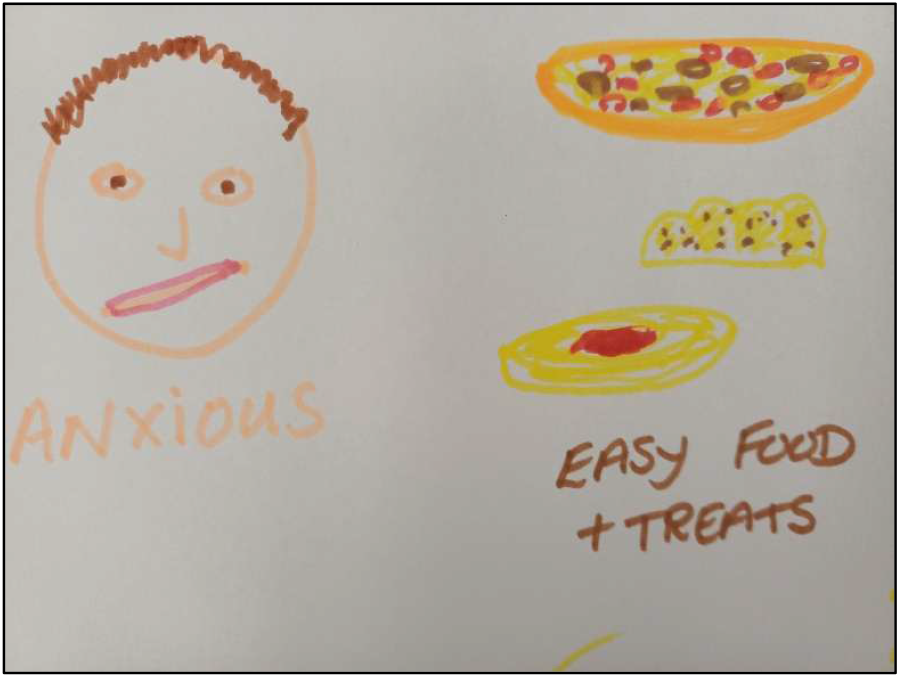
Storyboard extract, participant 026

Participants spoke of loss of control, particularly relating to portion sizes, which was often described as ‘self-sabotaging’ behaviour:

> *‘I’m happy. When I’m eating it. It’s only afterwards, once I’ve finished eating it, that actually I’m not any happier because of it, but in that moment I am. So that’s where I struggle with the self-sabotaging behaviour, I think*.*’* (participant 022)

Maintaining weight was almost universally perceived as formidable and laborious, particularly during the winter months, when participants believed they experienced an increase in negative thinking. Conversely, when participants felt more positive, they expressed greater feelings of self-control or were better able to adapt to keep on track:

> *‘it’s all about obviously getting your head in the right place as well, to then think right, long-term hopefully I can achieve my goals and lose the weight and reverse the damage I’ve done’* (participant 010)

Positive or negative thinking was a predictor for transition from a state of pre-intention to action for weight management. Interwoven through this was perception of body image. A desire for an idealistic body image and greater self-worth could motivate action. By contrast participant perceptions of their body image could induce feelings of shame, low self-esteem and feelings of judgement. Participants selected images or magazine word clippings relating to models in comparison to perceptions of themselves to express their perception of their own body image (Figure 4).

**Figure 4.**
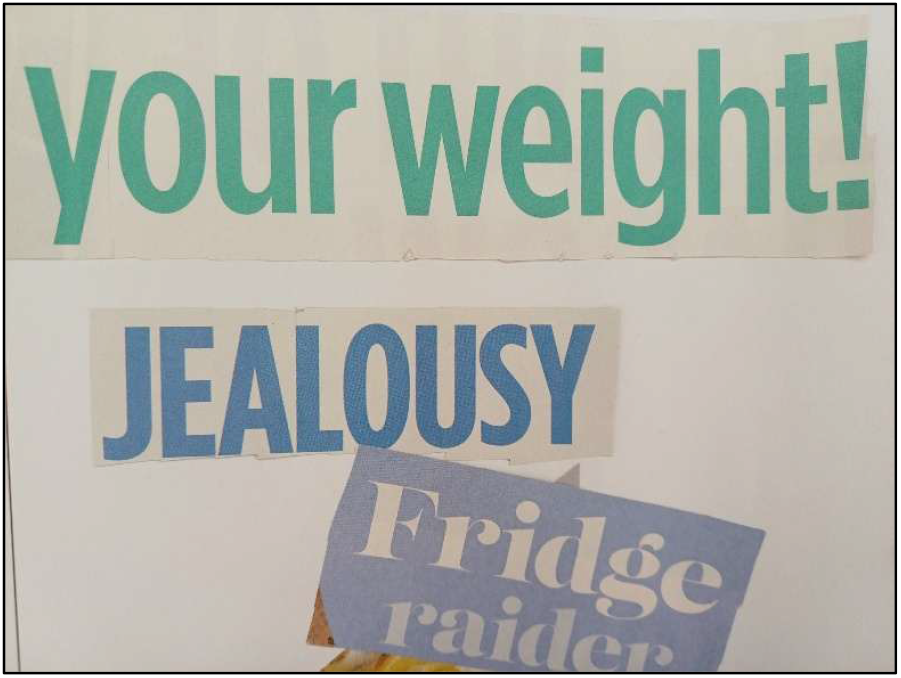
Storyboard extract, participant 007

Participant emotions appeared to be mediated by a range of internal and external factors. Key facilitators were receiving support for weight management, losing weight, feeling the benefits and experiencing higher perceptions of capabilities. Perceiving encouragement from social, peer and family groups was also closely related to feelings of confidence and success.

Participant capabilities appeared to be strongly related to having a more in-depth knowledge of nutrition, cooking skills, and how to undertake moderate activity which fitted into daily lifestyle. Greater exposure to facilitatory factors enabled the development of coping strategies and increased feelings of commitment to weight management:

> *‘you learn new things, tips, that kind of thing. You connect with your colleagues, your friends that are doing it, you feel boosted*.*’* (participant 028)

### ‘Yo-yo dieting’

A combination of contextual living circumstances and causal mechanisms of weight management influenced success for weight maintenance or perceived failure to maintain weight. A strong theme related to ‘yo-yo dieting’, a term participants applied to their constant cycling of weight changes. Participants perceived this as a vicious cycle which was frequently viewed as inescapable (Figure 5).

**Figure 5.**
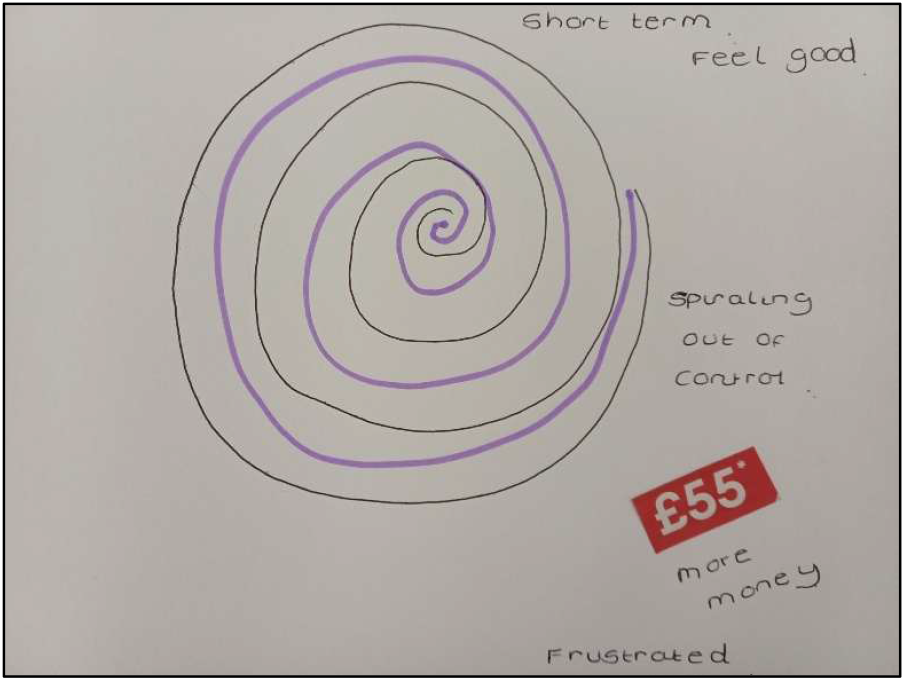
Storyboard extract, participant 030

Relapse to weight gain was frequently reported by participants and a commonly expressed experience of patient and public representatives also. A perceived increase in body size or breaking a goal resulted in feelings of failure, guilt and a lack of motivation to continue:

> *‘I do feel guilty if I’ve either put on weight or eaten something that I probably shouldn’t have done. Or if I’ve gone over my calories for a day or something, I’ll just feel really guilty and start comfort eating and then I’ve gone downhill again*.*’* (participant 014)

> *‘I get so I don’t care during. And motivational loss. And then I just think why bother? I know that’s really negative and I don’t mean to be negative but that’s just my journey with it*.*’* (participant 008)

Psychological mechanisms when compounded by contextual factors resulted in greater likelihood of relapse. Key contextual factors playing a role included cost of food, cost of support, and loss of support:

> *‘that was the 12-week free course from the council, but after that I couldn’t afford to keep doing it, so just sad that I couldn’t keep it going and seeing it through*.*’* (participant 009)

A minority of participants reported moving from ‘yo-yo’ dieting to a state of continued success of weight management. For these participants, success appeared to relate to taking small steps, making manageable changes which fitted with their lifestyle and that were sustainable over time:

> *‘no diet, no special diet, just eating sensibly, and just having little walks here and there, rather than use a car to go to the shops, walk up to the shop, things like that. That’s what helped me adjust and lose the weight*.*’* (participant 003)
>
> *‘I’m pretty disciplined on what I eat. I try to do … rounded sort of meals but I try to use healthy ingredients. And the other thing is instead of using a large plate, I use a smaller bowl so you can get less in it so you eat less*.*’* (participant 025)

### Weight management intervention needs

Participants expressed that ‘one size doesn’t fit all’ in terms of desired weight management support. There was agreement that ideally a service would offer options for both in-person and online or remote support and would offer both group sessions and one-to-one meetings.

Offering multiple options was thought important to ensure convenience and enable optimal fit with lifestyle. Participants also expressed that support should be affordable and easy to access and engage with. Participants were seeking support that included guidance for balanced flavourful eating, in addition to providing usable options for physical activity throughout the seasons.

There was a strong desire for longer-term support, or for a programme to include strategies that would boost individual capacities to maintain weight beyond an initial weight-loss programme. Two such strategies were improving knowledge and providing psychological support. Providing knowledge to extend understanding of weight management, was thought to enable greater long-term self-management of weight. Greater psychological support was also viewed as key to move from ‘yo-yo dieting’ to long-term weight maintenance.

Participants believed that there was a current lack of psychological support and a lack of support to address emotional eating, frequently perceived as the root cause of weight gain:

> *‘a lot of the issues behind eating is comfort and pleasure. And I don’t think that that’s being addressed*.*’* (participant 011)
>
> *‘someone might be struggling with their wellbeing or something in their background that needs to be addressed, that then they can focus on weight loss a little bit more and be healthier, ‘cause then they can break away from anything that’s impacting them’* (participant 034)

Participants believed that any programme should be respectful and encouraging in nature, to enable building of confidence, and to support making changes one step at a time. There was a need for holistic support, rather than a focus on change in body mass index; an index perceived as unhelpful. Tailored support to meet individual needs and preferences was expressed as the ideal.

Patient and public representatives collectively depicted the journey of moving from before, to during, and after an initial weight management programme. Their group storyboard reflected the same barriers and facilitators expressed by participants, and a universal desire for long term support and maintenance (Figure 6).

**Figure 6.**
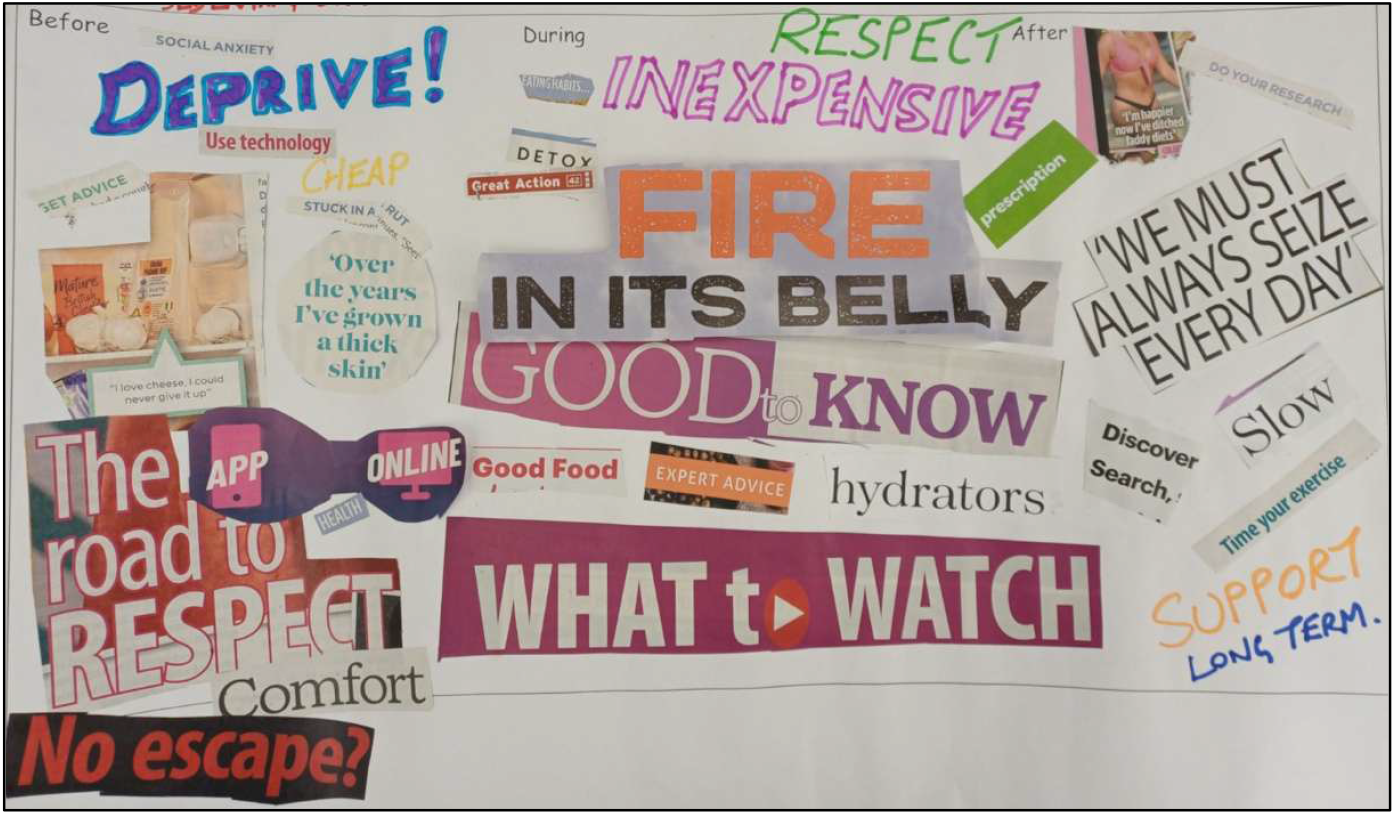
Patient and public representatives storyboard of weight management needs.

## Discussion

Storyboarding and application of a realist ‘lens’ were used to understand needs and experiences of weight management in under-resourced communities. This is the first study to use this combined approach to explore weight management needs. Key contextual barriers were cost and difficulties in accessing weight management services, healthy foods, and exercise opportunities. Key mechanisms acting as barriers to weight management were negative thinking and emotional eating, which resulted in cyclical ‘yo-yo dieting’.

Participants sought a weight management service that was flexible and that offered both in-person and remote support. For long-term weight maintenance, participants believed it was crucial to provide extended support and to include psychological support to address underlying causes of overeating. There was a desire for a service that was tailored to individual needs and medical circumstances.

Given the growing use of obesity medication [21], and evidence that weight regain is higher following cessation of medication compared to cessation of behavioural support [22], it is perhaps ever more important to identify which elements of behavioural weight management interventions are key for long-term weight maintenance, particularly within under-resourced communities. Our findings of key elements to include in a weight management programme add to and extend those in the literature. NICE guidance for overweight and obesity bases recommendations on the success of multicomponent interventions that are tailored to individuals and address long-term needs [23], themes which we also identified. Participants taking part in our research believed there was a lack of these interventions that were easy to access and of low-cost. Financial cost of programmes and transport limitations are recognised as barriers, particularly for low socioeconomic groups [24,25]. The inclusion of digital interventions may aid weight management within groups of lower socioeconomic status by providing an additional route of access [24]. Other barriers to accessing weight management support include health service factors such as low referral rates and inadequate communication with patients [7,25]. Perceived stigma of overweight and obesity, and limited support to address emotional eating also play significant roles [7,25]. To address stigma, we identified respect and encouragement to be important facilitators to encourage access and engagement with weight management services. We also found that participants perceived greater encouragement where small step changes with realistic goals were recommended. A review of behaviour changes techniques essential for weight loss also found goal setting, self-monitoring and commitment were important elements to include for success [6]. Participants were seeking increased knowledge and skills to address both nutrition and physical activity barriers, and to understand emotional eating. Addressing both nutrition and physical activity is recommended to increase effectiveness of weight management programmes [23]. Specific recommendations to include psychological therapies for supporting mental health in weight management programmes are lacking, in part due to lack of evidence for an effect [26].

However, there is growing recognition that psychological support, if added to traditional weight management programmes, would aid self-control of emotional eating [26,27].

### Study strengths and limitations

The strengths of this study lie in the storyboarding methodology which enabled an inclusive approach and expression from seldom heard voices. Allowing participants to express themselves by pictures or writing in addition to expressing themselves orally was valuable to collect in-depth views of the emotive topic of weight management. Viewing storyboards using a realist ‘lens’ enabled us to pinpoint key contextual and mechanistic factors that had an influence on weight management outcomes. This was a novel approach which provided a unique insight into experiences of overweight and weight management services. However, participants tended to present storyboards that focused on their personal journeys, rather than taking a wider view of weight management needs, which could be perceived as limiting generalisability [28]. Key findings were shared with participants and patient and public representatives. Participants requested no changes to results after viewing key findings. The views of patient and public representatives married well to the study findings which we believed increased validity of our results.

Approach of overweight individuals within community groups was challenging due to stigma of weight, eating disorders, privacy concerns and inclusivity. The criteria of ‘self-identifying’ as overweight was therefore emphasised, rather than a focus on body mass index, and community groups which focused on weight management or wellbeing were included. People who self-identify as overweight or who take part in weight management programmes, would likely more readily engage with weight management research, in comparison to the general population which may have reduced transferability of our findings. Nevertheless, we reached some participants who were not motivated to engage with weight management services. We also sought views of experiences before, during and after use of a weight management service to capture more information on the pre-contemplation phase. Reaching participants of a low socioeconomic status was also challenging. Whilst areas within the top 20% most deprived were targeted, participants did not all live within the targeted postcode areas.

Postcode by index of multiple deprivation was not stipulated as an inclusion criteria, since this would have been difficult to control for in community group settings. Reaching a socio-demographically diverse sample was also challenging, in part since the FMH Ethics Committee requested socio-demographics were collected following consent, which limited purposive sampling. Challenges of recruiting participants of lower socioeconomic status are widely recognised in the literature [29]. Irrespective of socioeconomic status, views of participants were broadly similar, with data saturation reached after six focus groups.

### Recommendations for further research

Further research is needed to test the success of interventions addressing the specific contextual and mechanistic factors that we have identified as key to weight management within under-resourced communities. Repeating this research with a more diverse sample, including underrepresented groups, would also be beneficial to test the generalisability of our findings.

## Conclusions

Our findings suggest that programmes to support weight management in under-resourced communities should include greater psychological support to address root causes behind weight gain, alongside advice or signposting to long-term support needs. Programmes should be low-cost and easy to access, and ideally tailored to individual needs and circumstances.

We recommend policy makers and practitioners incorporate these specific needs into weight management programmes to enable a move from ‘yo-yo’ dieting to long-term weight maintenance.

## Declarations

### Ethics approval and consent to participate

Ethical approval was granted by the FMH Research Ethics Committee at the University of East Anglia.

### Consent for publication

Participants provided oral and written consent to participate at the start of focus groups.

### Availability of data and materials

Data supporting the study findings are not publicly available due to privacy and ethical restrictions. Participants of this study did not agree for their data to be shared publicly.

### Authors’ contributions

Conceptualisation: ZK, TJB, KM, FN; Data curation: TJB, ZK; Formal analysis: TJB; Funding acquisition: ZK; Investigation: TJB, NAQT; Methodology: TJB, ZK, KM, FN; Supervision: ZK, FN; Writing-original draft: TJB; Writing-review and editing: All authors read and approved the final manuscript.

## Acknowledgements

We would like to sincerely thank our participants, patient and public representatives, and the following organisations and community groups: Feel Good Suffolk; Gainsborough Sports and Community Centre; Greenstead Community Centre; Healthwatch Essex; Ipswich Borough Council; Menscraft; Tendring District Council; The Feed; Whitton Sports and Community Centre.

