## Supplementary file 2 for "Weight management needs in under-resourced communities elicited using storyboarding and a realist lens: A qualitative study"

eSign activity sheet- experiencing existing weight-management services

**Creative thoughts**

*Think about your lifestyle e.g. money, friends/family, your health? How you would feel? What would help or hinder you using the service? Would you use the service? Would it help you manage your weight?*

| Before | During | After |
|---|---|---|

### **Written thoughts**

*Think about your lifestyle e.g. money, friends/family, your health? How you would feel? What would help or hinder you using the service? Would you use the service? Would it help you manage your weight*

Feelings:

Enablers:

Barriers:
